# A Pattern-Based Heart Rate Variability Approach in Somatic Symptom Disorder: Evidence from the SOMA.SSD Study

**DOI:** 10.1101/2025.11.25.25340969

**Authors:** Paul Hüsing, Wei-Lieh Huang, Kerstin Maehder, Franz Pauls, Yvonne Nestoriuc, Bernd Löwe, Kristina Blankenburg, Sophie Schmitz, Stefanie Hahn, Anne Toussaint

## Abstract

**Background:** Somatic Symptom Disorder (SSD) is characterised by a complex interplay between physiological regulation and psychological distress. Heart rate variability (HRV) has been proposed as a biomarker reflecting autonomic dysregulation in SSD, but single-parameter approaches have shown limited explanatory power. This study applied a proposed four-pattern HRV classification (normal, low, relatively high sympathetic, relatively high vagal pattern) to investigate its validity and its association with the SSD symptomatology in a German SSD sample. We examined whether these HRV patterns differentiate patients at baseline and predict trajectories of somatic and psychological symptoms over time.

**Methods:** Data from 148 German patients with SSD of the SOMA.SSD cohort were analyzed. HRV was measured at baseline via a 5-minute resting electrocardiogram (ECG) and classified into four patterns using age- and sex-adjusted European norms. Psychopathological outcomes (PHQ-15, PHQ-9, SSD-12) were assessed at baseline, 6, and 12 months. Between-group differences were analyzed using rmANOVA and linear mixed-effects models controlling for age and sex.

**Results:** Patients in the low HRV pattern group showed significantly higher levels of somatic symptom severity, depressive symptoms, and psychological distress compared to those with normal HRV pattern. No significant differences were observed among the high sympathetic and high vagal patterns. Across follow-ups, HRV pattern groups showed stable but distinct symptom levels with higher psychopathology scores in the low HRV pattern group, but without group × time interaction effects.

**Conclusions:** The four-pattern HRV approach was replicable in a Western SSD sample and identified a subgroup with low HRV and persistently elevated symptom burden. HRV-based pattern classification may serve as a feasible physiological marker for identifying autonomic subtypes in SSD and related disorders.

## 1. Introduction

Somatic Symptom Disorder (SSD) is a diagnosis first introduced in the Fifth Edition of the Diagnostic and Statistical Manual of Mental Disorders, (DSM-5)[1]. In contrast to the former ICD-10/DSM-IV category of somatoform disorders, SSD is characterized by distressing somatic symptoms accompanied by disproportionate thoughts, feelings, or behaviors. Prevalence rates are estimated between 4% and 5% within the adult German general population and much higher in specialized tertiary settings [2], and SSD is associated with increased functional impairment, decreased quality of life, and high comorbidity with anxiety and depressive disorders [3].

Given the emphasis on the interaction between physical symptoms and regulatory processes, increasing attention has been directed toward physiological correlates of SSD. One promising line of research has investigated heart rate variability (HRV) as a potential biomarker of SSD [4, 5]. HRV is widely regarded as a non-invasive measure of autonomic nervous system functioning [6], providing a variety of parameters that reflect sympathetic and parasympathetic regulation [7]. It has been used as an indicator for autonomic nervous system dysfunction in a variety of psychiatric disorders, including mood disorders, anxiety disorder, psychotic disorders, and substance abuse disorders [8–10]. Low HRV patterns have been linked to depressive symptoms, anxiety, and somatic symptom dimensions. Yet, the pronounced overlap and comorbidity among these symptom domains make it challenging to delineate the distinct contributions of autonomic dysfunction [11, 12]. Meta-analytic evidence indicates that individuals with SSD also tend to exhibit lower HRV compared to healthy controls [13]. However, the observed effects are typically of small-to-moderate magnitude and appear to be strongly moderated by comorbidities (e.g., depression, anxiety), as well as lifestyle and sociodemographic variables [4]. HRV measurement poses several methodological challenges [14], as it is known to be influenced by multiple sociodemographic and behavioural factors, such as age, sex, smoking habits, or medication intake (for example, betablocker or tricyclic antidepressants). Therefore, HRV assessments as well as data analysis should be controlled accordingly, if possible [15]. Although long-term (24-hour) recordings are often considered the gold standard for capturing the full spectrum of autonomic dynamics, however, they are time-intensive and thus often difficult to implement in research and clinical practice. Short-term resting-state recordings (e.g., 5-minute) are more feasible and widely used, and demonstrated acceptable reliability for several HRV parameters [7]. On the other hand, however, short-term recordings may capture only a limited snapshot of the overall autonomic functioning and might be more sensitive to situational influences. Despite these limitations, the feasibility of short-term recordings makes them particularly useful for diagnostic screening and subgroup classification in applied clinical settings [16]. However, relying on single HRV parameters only has thus far proven insufficient for diagnostic or prognostic purposes in patients with SSD. To address these limitations, Huang and colleagues (2022) proposed a pattern-based approach that classifies patients according to the constellations of four commonly assessed HRV parameters, using one time-based and three frequency-based parameters. Based on these indices and their corresponding normative data, Huang et al. distinguished four HRV patterns:

1. *Normal pattern* – characterized by all parameters within the expected normative range, indicating intact autonomic flexibility.
2. *Low pattern* – defined by globally reduced parameters, indicating an overall autonomic hypoactivity.
3. *Relatively high sympathetic*: parameters do not show a globally low tendency but a higher ratio of low-frequency to high-frequency power, indicating a balance slightly shifted toward sympathetic dominance.
4. *Relatively high vagal*: parameters do not show a globally low tendency but a lower ratio of low-frequency to high-frequency power, indicating a balance slightly shifted toward vagal dominance.

Patients undergoing resting-state HRV measurements can thus be assigned to one of the four HRV pattern. The authors based their classification on a large Korean reference sample [17] and used the standard procedure for 5-min HRV measurements, in which the participants were asked to sit and to breathe freely [16]. The measurement was in the daytime; the subjects were asked to rest for several minutes before the HRV recording. In their research, Huang and colleagues have used this HRV pattern categorization in a sample of patients with affective disorders and a control sample of healthy individuals and could show that a classification to the low HRV pattern was most strongly associated with depression, anxiety, and somatic symptoms, as well as age and body mass index (BMI). The relatively high sympathetic pattern and relatively high vagal pattern did not yield psychopathological differences from the normal HRV pattern, but were associated with personality traits and psychosocial factors, such as employment status and exercise habits [11]. Initial evidence overall suggests that the four HRV patterns may capture meaningful subgroups of patients beyond single-parameter approaches [18], and could thus offer important insights into factor contributing to SSD symptomatology. Given that HRV may be influenced by demographic, biological, and cultural factors [14], further validation and potential adaptation of the HRV patterns is needed. Since the HRV pattern structure has not yet been applied to longitudinal data, its predictive value regarding psychopathological development thus remains to be tested.

Against this background, the present study pursued three primary objectives. First, we aimed to replicate the proposed four-pattern HRV classification in a German sample of patients with SSD, using region-specific European normative data as a reference point. Second, we investigated whether stratifying patients according to these four HRV patterns would yield differences in sociodemographic factors and psychopathology at baseline, similar to the findings by Huang and colleagues [11]. Third, we examined whether distinguishing between the normal and low HRV pattern groups provides both exploratory power regarding the most common SSD-related psychopathology measures over time, namely somatic symptom severity, depression, and symptom-related psychological distress.

## 2 Methods

### 2.1 Participants and Procedure

Data for present study was derived from the SOMA.SSD study [19], which is part of the research unit “Persistent SOMAtic symptoms ACROSS diseases—from risk factors to modification” (RU SOMACROSS). This research unit, funded by the German Research Foundation (Deutsche Forschungsgemeinschaft, DFG), investigates biological and psychosocial processes underlying symptom persistence across different medical conditions [20]. Patient recruitment for the SOMA.SSD study was conducted at the outpatient clinic of the Department of Psychosomatic Medicine and Psychotherapy at the University Medical Center Hamburg-Eppendorf between August 2022 and November 2023. Baseline assessments (T0) and follow-up assessments at 6 months (T1) and 12 months (T2) involved both online questionnaires and study visits at the psychosomatic outpatient clinics. Inclusion criteria were age ≥18 years, a diagnosis of somatic symptom disorder (SSD) according to the German research version of the Structured Clinical Interview for DSM-5 [1], sufficient oral and written German language proficiency to complete self-report questionnaires and interviews. Exclusion criteria included serious illness requiring immediate intervention, florid psychosis or substance use disorder, and acute suicidality. The study was approved by the Ethics Committee of the Medical Association of Hamburg, Germany on 25 February 2021 (processing number: 2020-10196-BO-ff), and was conducted in accordance with the Declaration of Helsinki, Good Clinical Practice guidelines, and applicable national and local laws. Eligible patients received both verbal and written information prior to providing written informed consent.

### 2.2 Measures

Patients completed a comprehensive set of questionnaires at all assessment points (for a full overview, see [19]) and provided sociodemographic information relevant to HRV, including age, sex, smoking habits, and regular intake of prescribed medicine. Further assessments included measurement of anxiety (Generalized Anxiety Disorder Scale-7; GAD-7)[21], quality of life (Short Form Health Survey-12; SF-12)[22], and level of physical activity (International Physical Activity Questionnaire; IPAQ)[23]. Three outcomes were of special interest for this study: somatic symptom severity assessed using the Patient Health Questionnaire-15 (PHQ-15)[24], depressive symptom severity assessed using the Patient Health Questionnaire-9 (PHQ-9) [25], and the psychological distress due to the somatic complaints assessed with the Somatic Symptom Disorder ̶ B Criteria Scale (SSD-12)[26].

The Patient Health Questionnaire (PHQ) is a self-administered instrument derived from the PRIME-MD diagnostic tool that was designed to screen for common mental disorders [27]. The PHQ-15 is among the most widely used instruments for identifying individuals with an increased somatic symptom severity. It evaluates the presence and severity of common physical symptoms encountered in primary care — such as fatigue, gastrointestinal, musculoskeletal, pain-related, and cardiopulmonary complaints — over the preceding four weeks, using 15 items. Each symptom is rated on a three-point scale from 0 (“not bothered at all”) to 2 (“bothered a lot”). Total scores range from 0 to 30, representing the self-reported symptom severity, with higher scores indicating higher levels of symptom severity (0–4: none to minimal; 5–9: low; 10–14: moderate; 15–30: high). Internal consistency in common epidemiological samples ranges around Cronbach’s α = 0.80 [24]. The PHQ-15 has demonstrated sufficient to good psychometric properties and is recommended for the use in large-scale epidemiological and clinical studies [28]. The German adaptation likewise shows good? psychometric quality [29].

The PHQ-9 represents the depression subscale of the PHQ and assesses the nine depressive symptoms according to the DSM-IV, each rated from 0 to 3 [30]. Its good reliability and validity have already been confirmed in numerous international studies [25], and a well-validated German version is available [31].

The Somatic Symptom Disorder – B Criteria Scale (SSD-12) captures the psychological criteria of DSM-5 somatic symptom disorder [26]. It includes 12 items for four domains, each covering the three psychological subcomponents and rated from 0 to 4. The SSD-12 has shown good psychometric properties [32, 33]. Normative data have been established based on a large German cohort [33], thus enabling comparisons with representative population samples. The scale was originally developed in German [26].

### 2.3 Heart Rate Variability (HRV)

Resting-state HRV was assessed as part of the on-site study visit using the ambulatory sensors *Movisens ecgMove 4* (ECG sampling rate: 1024 Hz) with correction for potential artefacts in *DataAnalyzer [34]*. Measurement took place in the supine position for a duration of 6 minutes after a short resting phase prior to recording. HRV was then analyzed following established guidelines [14, 16] and artifacts in ECG recordings were identified and manually corrected. Both time-domain and frequency-domain HRV parameters were extracted for the purpose of this study. HRV patterns were specified based on the definition by Huang and colleagues [11] (table 1). It classifies patients according to constellations of four commonly assessed HRV parameters: standard deviation of normal–to–normal RR intervals (SDNN), low frequency power (LF), high frequency power (HF), and ratio of low-frequency to high-frequency power (LF/HF). SDNN reflects overall variability across the recording period and is regarded as an index of global autonomic flexibility. LF and HF are frequency-domain measures, with LF typically linked to a mixture of sympathetic and parasympathetic influences, and HF more directly associated with parasympathetic (vagal) activity. The LF/HF ratio has often been interpreted as an indicator of autonomic balance, although this interpretation remains debated [7].

**Table 1.** Definition of the four HRV patterns based on the recommendations by Huang et al. (2022)

| HRV patterns | Definition |
| --- | --- |
| Normal pattern | SDNN, LF and HF: at least two moderate to high<br>+ Moderate level of LF/HF |
| Low HRV pattern | SDNN, LF, HF: none or one moderate to high<br>+ Any level of LF/HF |
| Relatively high sympathetic pattern | SDNN, LF, HF: at least two moderate to high<br>+ High level of LF/HF |
| Relatively high vagal pattern | SDNN, LF, HF: at least two moderate to high<br>+ Low level of LF/HF |

*Note.* SDNN = standard deviation of normal-to-normal RR intervals; LF = low-frequency power; HF = high-frequency power; HRV = heart rate variability; LF/HF = ratio of low-frequency to high-frequency power

For our sample we referred to a recently published large Danish cross-sectional study [35] to define adjusted ranges of the moderate level, including separate norm values for different age groups and sex. In this study, 6891 participants aged 18–72 years were resting for 5 minutes before the measurement of continuous heart rate during 7 minutes of supine rest and normal breathing took place. In accordance with Huang and colleagues [11], the moderate level was defined as the range between the 25^th^ and 75^th^ percentiles.

### 2.4 Statistical analysis

Statistical analyses were performed using SPSS version 25 [36], with an α error probability set at 0.05 for statistical significance. After providing demographic characteristics and descriptive statistics for the overall sample, we compared sociodemographic and psychopathological variables across the four HRV pattern groups at baseline. For categorical sociodemographic variables (i.e., sex, medicine intake, smoking status), group differences were analyzed using Pearson’s chi-square tests of independence. Mean differences in continuous sociodemographic variables and psychopathological measures were examined using one-way ANOVA, followed by Tukey’s HSD post hoc tests where appropriate. In case homogeneity of variances was violated, we employed Welch ANOVA. To account for multiple testing, the Benjamini-Hochberg procedure was applied to control the false discovery rates in case of significant *p*-values. All tests were two-sided, and p-values below 0.05 after correction were considered statistically significant. Effect sizes for ANOVA were interpreted using conventional benchmarks for partial eta squared (ηp² = .01 small, .06 medium, .14 large)[37]. For longitudinal comparisons, only the normal HRV and low HRV pattern groups were used, as prior research suggests that only the low HRV pattern shows significantly lower levels of depression, anxiety, and somatic symptoms at baseline. Accordingly, linear mixed-effects models (LMMs) were fitted separately for PHQ-9, PHQ-15, and SSD-12. Each model included fixed effects of group (HRV normal pattern vs. HRV low pattern), time (T0, T1, T2), the group × time interaction, and the covariates age (in years) and sex (female, male). A random intercept accounting for within-subject correlation was included in the models and an unstructured covariance matrix was specified for residuals. Restricted maximum likelihood (REML) estimation was used and estimated marginal means were compared using Bonferroni adjustment. For all significant main and interaction effects in the linear mixed-effects models, partial eta squared (ηp²) values were calculated as measures of effect size.

## Results

### 3.1 Demographic and descriptive sample characteristics

The total SOMA.SSD sample consisted of *N* = 241 patients with SSD at baseline and HRV data were collected for *N* = 148 of those patients. Baseline characteristics of this sample are presented in Table 3. The majority of included patients could be assigned to either the normal or the low HRV pattern group (68.9 %), and the rest to the relatively high sympathetic (20.9 %) or the relatively high vagal pattern (10.1 %).

**Table 3:** Baseline characteristics of the total sample (*N* = 148)

| Variable | Mean (*SD*) or n (%) |
| --- | --- |
| Age (years) | 45.1 (12.8) |
| Sex |  |
| *female* | 97 (65.5 %) |
| *male* | 51 (34.5 %) |
| Marital status |  |
| *single* | 81 (54.7 %) |
| *married* | 48 (32.4 %) |
| *other* | 19 (12.9 %) |
| Educational status |  |
| < 10 years of school | 12 (8.1 %) |
| 10 years of school | 28 (18.9 %) |
| 11 - 13 years of school ("Abitur") | 57 (38.5 %) |
| Higher education / University degree | 47 (31.8 %) |
| *Other* | 2 (1.4 %) |
| Employment status |  |
| *Employed* | 95 (64.2 %) |
| *Unemployed* | 19 (12.8 %) |
| *Not in the work force* | 34 (23.0 %) |
| Current intake of prescribed medicine |  |
| *yes* | 110 (74.3 %) |
| *no* | 38 (25.7 %) |
| Smoking |  |
| *yes* | 29 (19.6 %) |
| *no* | 119 (80.4 %) |
| HRV group pattern |  |
| *Normal* | 61 (41.2 %) |
| *Low* | 41 (27.7 %) |
| *Relatively high sympathetic pattern* | 31 (20.9 %) |
| *Relatively high vagal pattern* | 15 (10.1 %) |

Note. HRV = Heart Rate Variability. Normal HRV group pattern = SDNN, LF, HF at least two moderate to high + moderate level of LF/HF; low HRV group pattern = SDNN, LF, HF none or one moderate to high + any level of LF/HF; Relatively high sympathetic pattern = SDNN, LF, HF at least two moderate to high + high level of LF/HF; Relatively high vagal pattern = SDNN, LF, HF at least two moderate to high + low level of LF/HF

### 3.2 HRV patterns and differences at baseline

Regarding sociodemographic variables, significant differences in the frequency distributions among the HRV pattern groups were found regarding intake of prescribed medication (χ²(3) = 12.905, p = .005, φ = .295), with more patients in the low HRV pattern group taking medication (95.1%) compared to all other groups (normal pattern 65.6 %, high sympathetic pattern 67.7 %, high vagal pattern 66.7%).

Regarding psychopathological variables, we found slightly higher levels of somatic symptom severity, depressive symptoms, and psychological distress due to the somatic symptoms in patients in the low HRV pattern group, but also lower levels of physical quality of life and activity levels compared to all other HRV pattern groups. However, these differences were not found to be statistically significant (Table 4). Post-hoc analysis just indicated a significant difference (*p* = 0.047) in physical quality of life between the normal and low HRV pattern groups (mean difference = 5.48, 95 %-CI[0.05, 10.9]).

**Table 4.** Descriptive statistics for sociodemographic variables and psychopathology across the HRV pattern groups at baseline including group comparisons.

| Variable | Normal pattern<br>(n = 61) | Low pattern (n= 41) | High sympathetic<br>pattern (n = 31) | High vagal<br>pattern (n = 15) | Group comparison |
| --- | --- | --- | --- | --- | --- |
|  | Mean (SD) | Mean (SD) | Mean (SD) | Mean (SD) | Statistics |
| *Sociodemographics* |  |  |  |  |  |
| Age | 47.3 (13.12) | 44.1 (12.24) | 41.2 (12.43) | 46.6 (12.82) | $F(3, 144) = 1.752, p = 0.159$ |
| Sex, female | 42 (68.9 %) | 23 (56.1 %) | 23 (74.2 %) | 9 (60.0 %) | $\chi^2(3) = 3.147, p = 0.370$ |
| **Intake prescribed medicine, yes** | **40 (65.6 %)** | **39 (95.1 %)** | **21 (67.7 %)** | **10 (66.7 %)** | **$\chi^2(3) = 12.905, p = 0.005$** |
| Smoking, yes | 14 (23.0 %) | 7 (17.1 %) | 5 (16.1 %) | 3 (20.0 %) | $\chi^2(3) = 0.839, p = 0.840$ |
| *Psychopathology* |  |  |  |  |  |
| Somatic Symptom Severity (PHQ-15) | 11.4 (4.83) | 13.5 (5.1) | 12.2 (5.05) | 11.3 (5.32) | $F(3, 144) = 1.644, p = 0.182$ |
| Depression Severity (PHQ-9) | 11.5 (4.82) | 13.4 (6.28) | 11.4 (4.18) | 12.8 (5.02) | $F(3, 51.49) = 1.213, p = 0.314$ |
| Psychological Distress due to Somatic Symptoms (SSD-12) | 26.4 (8.97) | 29.9 (9.5) | 28.6 (9.13) | 29.3 (6.96) | $F(3, 144) = 1.366, p = 0.256$ |
| Anxiety severity (GAD-7) | 10.2 (4.8) | 10.7 (5.79) | 10.5 (4.72) | 11.4 (5.05) | $F(3, 144) = 0.235, p = 0.872$ |
| Physical Quality of Life (SF-12-P) | 39.9 (11.13) | 34.4 (9.14) | 38.7 (10.36) | 36.6 (9.97) | $F(3, 144) = 2.437, p = 0.067$ |
| Mental Quality of Life (SF-12-M) | 31.8 (10.1) | 36.0 (12.8) | 32.6 (9.82) | 34.1 (8.83) | $F(3, 53.17) = 1.083, p = 0.364$ |
| Activity level (IPAQ), at least moderate | 43 (72.9 %) | 23 (56.1 %) | 26 (83.9 %) | 10 (66.7 %) | $\chi^2(3) = 6.907, p = 0.075$ |

**Note.**; PHQ = Patient Health Questionnaire, GAD-7 = General Anxiety Disorder Scale-7; SSD-12 = Somatic Symptom Disorder – B-Criteria Scale-12; SF-12 = Short Form Health Survey-12; IPAQ = International Physical Activity Questionnaire. Normal HRV group pattern = SDNN, LF, HF at least two moderate to high + moderate level of LF/HF; low HRV group pattern = SDNN, LF, HF none or one moderate to high + any level of LF/HF; Relatively high sympathetic pattern = SDNN, LF, HF at least two moderate to high + high level of LF/HF; Relatively high vagal pattern = SDNN, LF, HF at least two moderate to high + low level of LF/HF.

### 3.3. Group (normal vs. low HRV pattern) x time effects on psychopathology

At baseline, *n* = 61 patients with SSD were classified to the HRV normal group and *n* = 41 patients to the HRV low group. Trajectories for the mean levels of somatic symptom severity (PHQ-15; Figure 1), depressive symptoms (PHQ-9; Figure 2) and psychological burden due to somatic symptoms (SSD-12; Figure 3) are depicted for each HRV pattern group.

**Figure 1.**
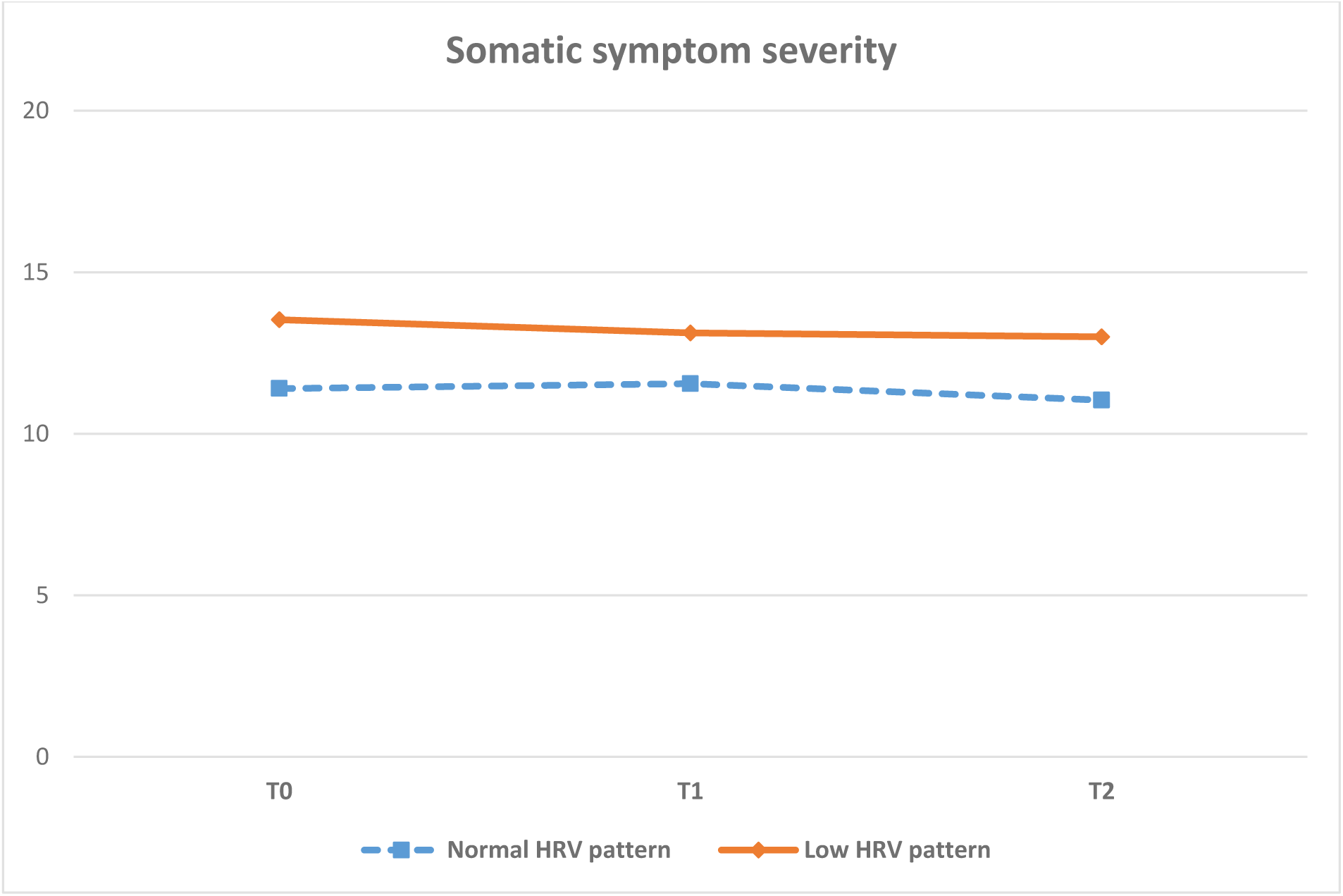
Sum scores of somatic symptom severity (PHQ-15) for normal HRV and low HRV pattern group at baseline (T0), after 6 months (T1), and after 12 months (T2).

**Figure 2.**
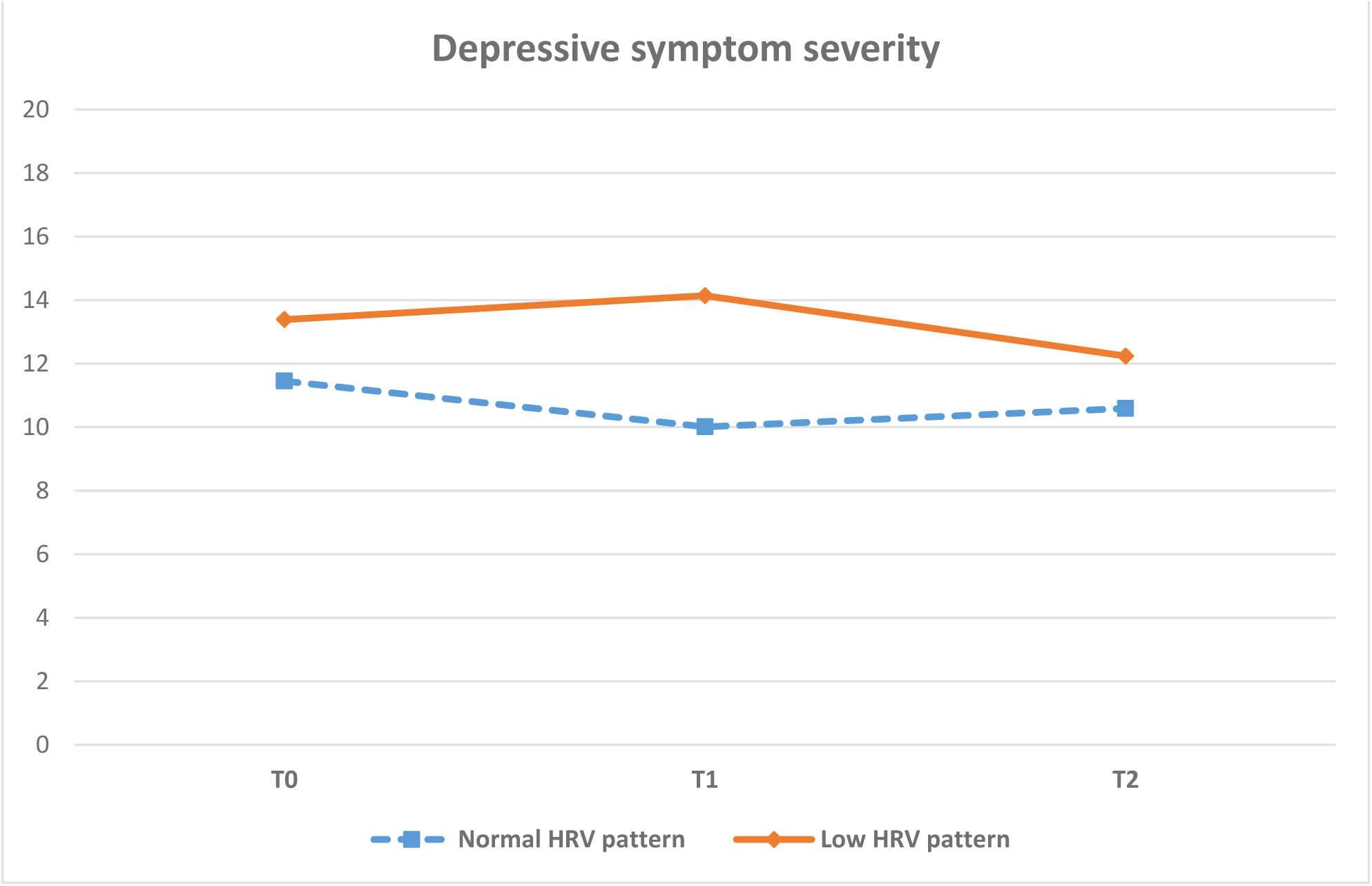
Sum scores of depressive symptoms (PHQ-9) for normal HRV and low HRV pattern group at baseline (T0), after 6 months (T1), and after 12 months (T2).

**Figure 3.**
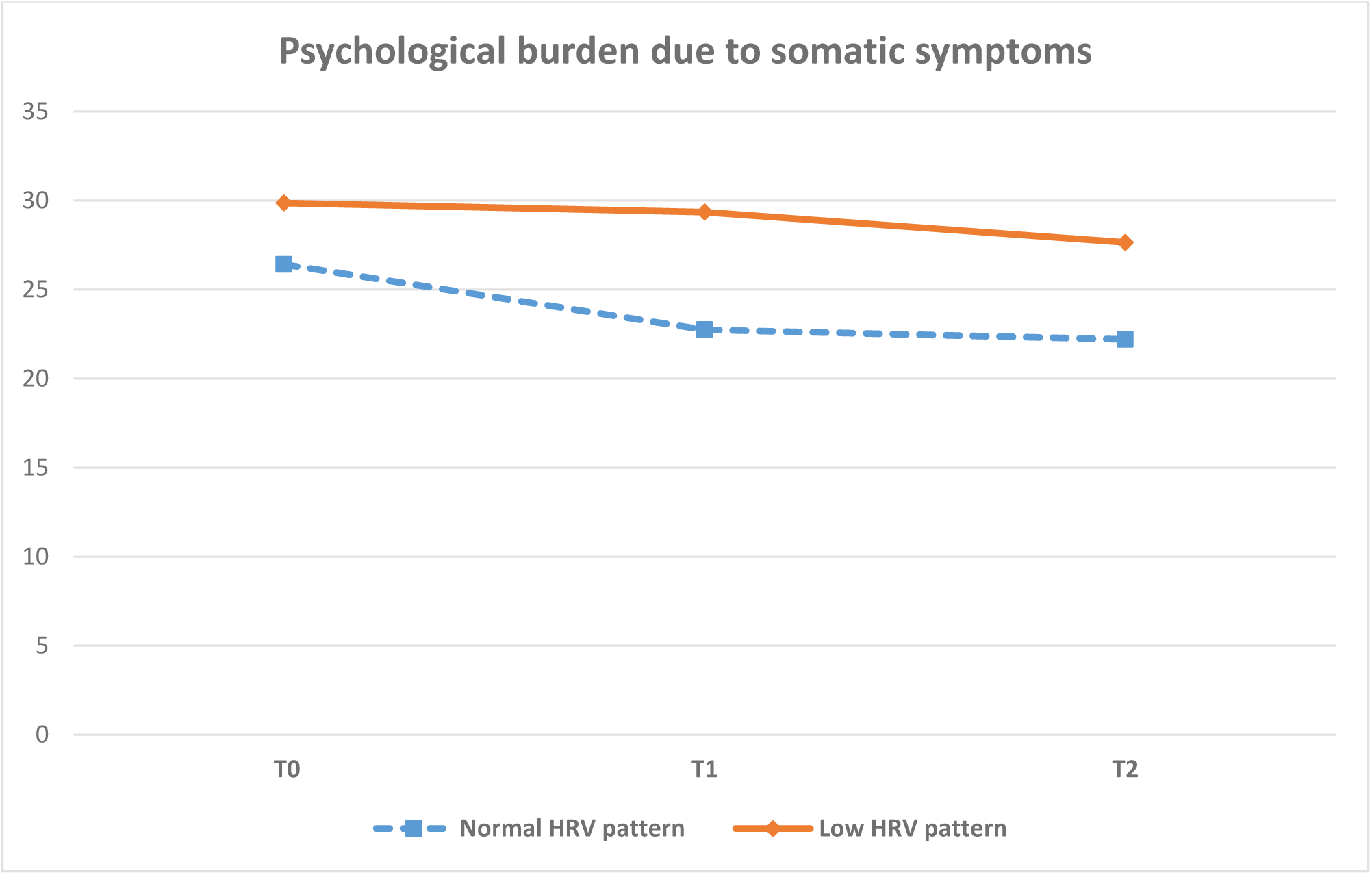
Sum scores of psychological burden due to somatic symptoms (SSD-12) for normal HRV and low HRV pattern group at baseline (T0), after 6 months (T1), and after 12 months (T2).

For all three criterion variables (PHQ-15, PHQ-9, and SSD-12), linear mixed-effects models revealed significant small-to-medium main effects of group (HRV pattern group), with participants in the low HRV pattern group reporting higher levels of symptom severity, PHQ-15, *F*(1, 276) = 11.52, *p* < 0.001, *ηp²* = 0.04, depressive symptom severity, PHQ-9, *F*(1, 270) = 14.59, *p* < 0.001, *ηp²* = 0.05, and psychological distress due to somatic symptoms, SSD-12, *F*(1, 269) = 18.45, *p* < 0.001, *ηp²* = 0.07, than those in the normal HRV pattern group. For the PHQ-15, there was also a significant medium-sized main effect of sex, F(1, 276) = 15.32, *p* < .001, *ηp²* = 0.053, with women reporting higher levels of symptom severity than men. No significant main effects of time or group × time interactions were observed, indicating that scores remained stable across measurement points, with consistent group differences (and sex differences for PHQ-15).

## 3. Discussion

The present study replicated the four-pattern approach of resting-state heart rate variability (HRV), originally proposed by Huang et al. (2022), in a German sample of patients with somatic symptom disorder (SSD) using European normative data. We also investigated whether these HRV patterns may appropriately differentiate between patients according to their psychopathological severity and were associated with symptom change over 12 months, thereby expanding upon prior findings. Specifically, patients classified with a low HRV pattern consistently exhibited higher levels of somatic and depressive symptom severity, as well as greater psychological distress related to somatic symptoms, compared with those with a normal HRV pattern over time.

Our findings support the feasibility of applying the four-pattern HRV classification to a European clinical sample. The distribution of patients with SSD within the HRV pattern groups and associations between HRV patterns and psychopathological variables largely mirrored those reported in Asian populations. These results are in line with previous evidence linking reduced HRV to heightened emotional and somatic symptomatology [4, 5, 13] and reinforcing the interpretation of low HRV as an indicator of reduced autonomic flexibility and stress-regulation capacity. Psychopathological outcomes for the “high sympathetic” and “high vagal” HRV pattern groups did not differ meaningfully from the other HRV pattern groups. These patterns may instead reflect individual differences related to lifestyle factors or other factors, or personality traits rather than direct markers of symptom severity, as previously discussed by Huang et al. (2022). Future research should thus focus on the psychological and behavioral correlates of these subtypes, possibly by integrating personality, somatic illness and medication, stress reactivity, or activity-level measures.

From a clinical perspective, distinguishing between low and normal HRV patterns may help identify patients at risk for greater symptom burden and functional impairment using a biomarker rather than psychological measures, i.e. self-report. Relying on a single HRV parameter only is not recommended, as it may not capture the complex interplay of the autonomic nervous system and can be heavily influenced by factors such as recording length, body position, and external conditions [38, 39]. Although HF is generally regarded as a specific indicator of parasympathetic activity, when an individual’s breathing rate is low, however, LF or SDNN are often suggested to capture parasympathetic effects more reliably. In research where breathing rate is often unstandardized, considering the overall trends of HF, LF, and SDNN together may provide a more complete and reliable picture of an individual’s parasympathetic characteristics [18]. In contrast, single parameters may rather provide an incomplete picture of autonomic functioning and are likely prone to random error, making them unreliable for detecting small changes or assessing interventions without rigorous standardization. Because the required HRV parameters are already collected in standard short-term recordings, the four-pattern classification can be easily integrated into ongoing clinical assessments without additional instrumentation or cost. On the long term, this approach may also facilitate the development of biopsychophysiological screening tools for SSD and related disorders.

However, several limitations of the present study should also be acknowledged. First of all, HRV was measured only once under resting-state conditions and in the supine position, limiting insight into dynamic autonomic responses or adaptability to stress. Accordingly, the reference data used in this study also only applies to HRV measurements taken in the supine position and cannot be transferred to other measurement set-ups (i.e. measurements in a sitting position). Although the initial study design included follow-up HRV assessments, technical constraints and dropout rates prevented sufficient HRV sample sizes for these analyses. Given that HRV is sensitive to situational and state-dependent factors, future studies should incorporate repeated or ambulatory assessments to capture variability across contexts. Medication use might also represent a confounding factor, as most patients reported medication intake, which may affect HRV through pharmacological effects and corresponding somatic illness. Therefore, a more comprehensive stratification by medication type and dosage is therefore warranted. Despite these limitations, the pattern-based HRV approach shows promise for subgrouping patients with SSD along autonomic dimensions. Further validation is needed with longitudinal HRV endpoints and systematic control of sociodemographic and pharmacological confounders. Beyond its immediate clinical utility, clarifying the biological underpinnings of SSD via HRV could inform tailored interventions such as HRV biofeedback [40] and mechanistic research into the autonomic correlates of persistent somatic symptoms.

In summary, the present findings demonstrate the applicability and clinical potential of a pattern-based HRV approach in SSD. Low HRV appears to characterize a subgroup of patients with SSD that is likely to suffer from higher levels of somatic and psychological symptom burden, consistent across time. Future research should extend these results by incorporating dynamic HRV indices, controlling for pharmacological influences, and examining how autonomic profiles interact with psychological and behavioral treatment responses.

## Conflicts of interest

None to declare.

## Financial support

This study was carried out within the framework of Research Unit 5211 (FOR 5211) ’Persistent SOMAtic Symptoms ACROSS Diseases: From Risk Factors to Modification (SOMACROSS)’ 1, funded by the German Research Foundation (Deutsche Forschungsgemeinschaft, DFG). The DFG grant numbers for the present study are NE 1635/3-1 (YN) and TO 908/2-1 (AT), see also https://gepris.dfg.de/gepris/projekt/460374026. The funding agency was not involved in the design of this study and was not involved in implementation, analyses and interpretation of data, or decision to publish results.

## Acknowledgment

We would like to thank all participating patients for investing their time and efforts to help with our study.

## Statement of Ethics

All study procedures were approved by the ethics committee of the Medical Chambers Hamburg, Germany (reference number: 2020-10197-BO-ff, 25 January 2021). Participants gave informed consent to participate in the study before taking part.

## Declaration of Generative AI and AI-assisted technologies

During the preparation of this work, the authors used ChatGPT for spellchecking and to improve grammar. After using this tool, the authors reviewed and edited the content as needed and take full responsibility for the content of the publication.

## Data Availability Statement

Data are available upon reasonable request.

## Notes

### Competing Interest Statement

The authors have declared no competing interest.

### Clinical Trial

ISRCTN36251388

